# Association of Genetic Liability to Psychiatric Disorders with Peripheral Metabolic Dysregulation

**DOI:** 10.64898/2026.06.06.26354927

**Authors:** Juan F. De la Hoz, Younga Heather Lee, Justin D. Tubbs, William Meyerson, Mihael Cudic, Devon Watts, Yen-Chen Anne Feng, Yulu Chen, Jessica Lasky-Su, Tian Ge, Jordan W Smoller

**Affiliations:** Psychiatric and Neurodevelopment Unit, Center for Genomic Medicine, Massachusetts General Hospital, Boston, MA; Center for Precision Psychiatry, Department of Psychiatry, Massachusetts General Hospital, Boston, MA; Department of Psychiatry, Harvard Medical School, Boston, MA; Stanley Center for Psychiatric Research, Broad Institute of MIT and Harvard, Cambridge, MA; Broad Trauma Initiative, Broad Institute of MIT and Harvard, Cambridge, MA; Institute of Health Data Analytics and Statistics, College of Public Health, National Taiwan University, Taipei, Taiwan; Channing Division of Network Medicine, Brigham and Women’s Hospital, Boston, MA

## Abstract

**Importance:** Individuals with psychiatric disorders face elevated cardiometabolic risk which is linked to increased mortality. The extent to which this reflects shared pathogenesis or the downstream effects of illness and treatment remains poorly understood.

**Objective:** To characterize the direct pleiotropic effects of psychiatric genetic liability on circulating metabolites and aggregate cardiometabolic risk, independent of psychiatric diagnosis and psychotropic medication use.

**Design, Setting and Participants:** Cross-sectional analysis of Mass General Brigham Biobank participants with metabolomic profiling, genomic data, and linked electronic health records.

**Exposures:** Genetic liability to nine psychiatric disorders quantified using polygenic risk scores (PRS): attention deficit/hyperactivity disorder (ADHD), anorexia nervosa (ANO), anxiety disorder (ANX), autism spectrum disorder (ASD), bipolar disorder (BD), major depressive disorder (MDD), PTSD, schizophrenia (SCZ), and substance use disorder (SUD).

**Main Outcomes and Measures:** 249 circulating metabolites and four metabolomic risk scores (MRS) for type 2 diabetes, myocardial infarction, ischemic stroke, and vascular dementia. PRS-metabolite associations were estimated using nested models adjusting for lifetime psychiatric diagnosis and psychotropic medication use.

**Results:** Across 25,290 participants, we identified 604 significant PRS-metabolite associations (Bonferroni p< 1.36 × 10^-4^), of which 89% persisted after adjustment for lifetime diagnosis and medication use, suggesting that the direct genetic effects on metabolism are largely independent of illness or treatment. PRS for MDD, PTSD, and ADHD showed the most extensive dysregulation, with a transdiagnostic pattern of elevated lipids and systemic inflammation, specifically triglycerides (β = 0.04 to 0.05, all p< 4.4 ×10^-13^) and glycoprotein acetyls (β = 0.05, all p< 2.2 ×10^-16^). Notably, PRS for SCZ and BD showed minimal metabolite dysregulation despite having the strongest association with their target diagnoses. PRS for MDD, PTSD, ADHD, and SUD were associated with increased MRS across cardiometabolic conditions (β = 0.03 to 0.08, all p< 2.1 ×10^-4^). Sensitivity analyses controlling for BMI or excluding participants without any psychiatric history (N: 21,305 and 11,150, respectively) showed a similar pattern.

**Conclusions and Relevance:** Psychiatric genetic liability is associated with systemic metabolic dysregulation independent of illness onset or treatment, supporting a partially pleiotropic basis for psychiatric-cardiometabolic comorbidity.

**Key findings:** *Question:* Does genetic liability to psychiatric disorders influence peripheral metabolism independent of psychiatric diagnosis and treatment?

*Findings:* In 25,290 participants, polygenic scores for major depressive disorder, posttraumatic stress disorder, and attention deficit hyperactivity disorder showed widespread associations with metabolic dysregulation, such as elevated triglycerides and systemic inflammation, which persisted after adjusting for diagnosis and medication use. Polygenic scores for schizophrenia and bipolar disorder showed minimal metabolite associations despite their well-established cardiometabolic burden.

*Meaning:* Shared genetic pathways partially underlie psychiatric and cardiometabolic comorbidity, implicating transdiagnostic biomarker profiles.

## Introduction

Psychiatric disorders are associated with elevated rates of cardiometabolic disease, leading to reduced quality of life, increased healthcare costs, and premature mortality ^1^. This association spans different conditions: individuals with depression are at increased risk for type 2 diabetes (T2D) and dyslipidemia ^2^, individuals with ADHD have an elevated risk of hypertension and T2D in adulthood ^3^, and those with schizophrenia have more than twice the rates of metabolic syndrome (MetS) as the general population ^4^. The etiology of this comorbidity is likely multifactorial ^5^, including adverse metabolic effects of pharmacological treatment ^6^, symptoms and behaviors associated with psychiatric conditions (e.g. sleep disturbance, altered diet, smoking) ^7^, and shared genetic factors underlying psychiatric and cardiometabolic conditions ^8^. A detailed understanding of their common pathophysiology could inform the development of integrated treatment strategies that simultaneously address psychiatric and cardiometabolic risk.

Metabolomics, the large-scale profiling of small molecules in blood, offers an opportunity to identify biological pathways linking psychiatric and cardiometabolic disease. Case-control studies of individual psychiatric disorders have documented altered metabolic profiles across multiple disorders, including PTSD ^9^, depression ^10^, and ADHD ^11^. However, these studies have not isolated the effects of genetic liability from those of illness severity and pharmacological treatment, all of which are confounded in affected individuals, and have largely focused on individual disorders rather than spanning the broader spectrum of psychiatric disorders ^12^. Polygenic risk scores (PRS) provide a quantitative measure of this inherited liability that precedes illness onset, providing an opportunity to estimate the effect of genetic risk for psychiatric disorders on metabolism. While previous work has done this for cardiometabolic disease ^13^, the specific metabolites underlying these associations have not been systematically characterized, and whether they reflect direct genetic effects independent of psychiatric illness and treatment has not been evaluated.

Here, we quantify the associations between polygenic liability for nine psychiatric disorders and 249 circulating metabolites as well as four composite metabolic risk scores for cardiometabolic outcomes in participants of a large biobank with linked electronic health records (EHR). To isolate direct genetic effects on peripheral metabolism, we test whether these associations persist after adjusting for psychiatric diagnosis and psychotropic medication use.

## Methods

### Study Population and Cohort Definition

The Mass General Brigham Biobank (MGBB) is a research program launched in 2010 that links genomic data, blood-based biomarkers, and longitudinal EHR for adult patients receiving care at MGB-affiliated hospitals and outpatient clinics ^14,15^. Participants provide informed consent for broad research use and donate blood samples for plasma, serum, and DNA extraction. Further details on participant recruitment and the consent process can be found in a prior publication ^14^.

### Construction of Polygenic Risk Scores

The biobank has generated genotyping microarray data using the Illumina Multi-Ethnic Genotyping Array (MEGA) and Global Screening Array (GSA) platforms. We performed standard quality control procedures and imputation using the Haplotype Reference Consortium Panel (eMethods). We generated PRS for nine psychiatric disorders (schizophrenia, SCZ; bipolar disorder, BD; major depressive disorder, MDD; anxiety disorders, ANX; post-traumatic stress disorder, PTSD; attention deficit/hyperactivity disorder, ADHD; substance use disorder, SUD; autism spectrum disorder, ASD; and anorexia nervosa, AN) using summary statistics from the largest available GWAS (eTable 1) and PRS-CS-auto ^16^, a Bayesian method that infers optimal shrinkage parameters from the data without requiring a validation sample for hyperparameter tuning. With these SNP weights, we used PLINK2 ^17^ to calculate PRS among an unrelated subset of MGBB participants. We restricted our sample to participants with European-like ancestry, reflecting both the predominant composition (∼85%) of MGBB, and the limited transferability of PRS derived from European-like GWAS summary statistics to other ancestry groups.

### Psychiatric Phenotype Ascertainment

We used participants’ clinical records to derive two sets of binary mediator variables: nine diagnosis variables (one per psychiatric disorder), and five psychotropic medication variables (one per medication class: see below). For each disorder, we mapped codes from the International Classification of Diseases to phecodes using the PheWAS R package ^18^, with the full list of corresponding codes provided in eTable 2. A participant was coded 1 if at least one corresponding phecode appeared in their EHR record, and 0 otherwise. We used a threshold of one phecode to maximize sample size. In sensitivity analyses, we required at least two phecodes for defining cases, which has been shown to improve positive predictive value ^19^. We used complete EHR records rather than restricting to diagnoses recorded before the blood draw, as diagnostic delay can result in years between true illness onset and the first recorded code ^20^. We classified psychotropic medications into five classes ^6^: antidepressants (Ad), antipsychotics (Ap), mood stabilizers (Ms), stimulants (St), and benzodiazepines (Bz) (eTable 3). Unlike with diagnoses, where we used lifetime history, we required at least two prescriptions within the year preceding the blood draw, as we aimed to capture medication use around the time of metabolomic profiling.

### Nightingale Metabolomic Panel

Following previously published protocols ^21^, Nightingale Health Plc. used nuclear magnetic resonance (NMR) spectroscopy to profile plasma samples collected during routine clinical blood draws (between 2010-2023). The Nightingale panel consists of 249 metabolite measurements spanning multiple classes, including lipoprotein particle concentrations, lipid constituents, fatty acids, amino acids, glycolysis-related metabolites, ketone bodies, and inflammation markers (eTable 4). We preprocessed and quality controlled raw values for each metabolite following established protocols ^22^, and excluded participants with 5 or more missing values (eMethods).

### Construction of Metabolomic Risk Scores

Following the approach of Barrett et al. ^23^, we constructed patient-specific metabolomic risk scores (MRS), which aggregate metabolite values weighted by their association with incident disease risk, adjusted by age and sex. We generated MRS for four conditions: type 2 diabetes (T2D), myocardial infarction (MI), ischemic stroke (IS), and vascular and other dementia (VOD). Each score was calculated as follows:

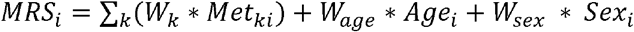

where *Met*_*ki*_ is the value of metabolite *k* for individual *i*, and where *W* are the published weights from ^23^, derived from Cox models linking metabolite levels to time to incident disease in UK Biobank participants.

### Statistical Analysis

Within the analytic sample, we standardized the PRS, MRS and metabolite values to a mean of 0 and standard deviation of 1. To evaluate the performance of each PRS, we tested their association with the corresponding psychiatric diagnosis using logistic regression, adjusting for age, chromosomal sex, and the first ten genetic principal components. We estimated the variance explained (R^2^) on the liability scale using the method of Lee et al. ^24^, meta-analyzing estimates across GSA and MEGA chips. Assumed population prevalences are provided in eTable 1.

To isolate direct genetic effects on metabolism from those operating through illness and treatment, we fit nested linear regression models for each PRS-metabolite pair. The *total model* estimated the effect of a psychiatric PRS on a metabolite, adjusting only for age at blood draw, sex, and the first ten genetic principal components, and the *direct model* additionally adjusted for the lifetime history of the respective psychiatric diagnosis and use of any psychotropic medication in the preceding year.

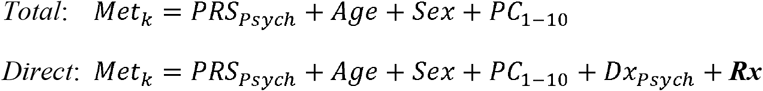

where *PRS*_*Psych*_ is the PRS for a given psychiatric disorder, *Dx*_*Psych*_ is the binary diagnosis variable for that same disorder, and ***Rx*** is a vector of five binary medication variables. All five medication classes were included regardless of the PRS being tested, to reflect the transdiagnostic nature of psychotropic prescribing ^25,26^. We applied the same nested modeling framework to test associations between each PRS-MRS pair, with *MRS*_*CDM*_ replacing *Met*_*k*_ as the outcome variable, where *CDM* denotes each of four cardiometabolic conditions (T2D, MI, IS, and VOD).

To characterize how much of each PRS-metabolite association reflects a direct genetic effect independent of diagnosis and medication, we computed the percent change in the beta coefficient as:

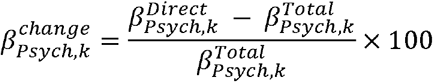

where 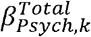 and 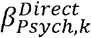 are the estimated coefficients for *PRS*_*Psych*_ on *Met*_*k*_ in the *total* and *direct effect* models, respectively. We also report the proportion of significant associations in the *total effect model* that remained significant in the *direct effect model*.

All models were fitted separately for participants genotyped on GSA and MEGA chips, and effect estimates were combined using fixed-effects inverse-variance weighted meta-analysis. To correct for multiple testing, we applied Bonferroni correction based on 41 independent metabolite principal components ^27^ and 9 PRS, yielding 369 tests and a significance threshold of p < 1.36 ×10^-4^ (0.05 / 369). For MRS analyses, we applied Bonferroni correction across 36 tests (9 PRS × 4 MRS), yielding a significance threshold of p < 1.38 ×10^-3^ (0.05 / 36). All analyses were conducted in Python (v3.13.1) and R (v4.5.2).

We conducted two additional sensitivity analyses. First, to assess the robustness of PRS-metabolite associations to residual confounding by psychiatric comorbidity, we restricted the sample to participants with no documented history of any psychiatric diagnosis or psychotropic medication use and fit the total model in this subsample. Second, body mass index (BMI) is strongly associated with circulating metabolite levels, but whether it acts as a mediator or collider of PRS-metabolite associations is uncertain. Among participants with BMI measured within one year of the blood draw, we averaged all available BMI measurements within that window and fit the direct model with additional adjustment for mean BMI.

### Ethical approval

This research was conducted as part of the “Mental Health and Chronic Disease: A PsycheMERGE Investigation into the Shared Biology Underlying Psychiatric Disorders” Study, with approval from the MGB Institutional Review Board (IRB Protocol ID: 2019P003696).

## Results

### Sample characteristics and PRS validation

Our sample consisted of 25,290 MGB Biobank participants with metabolomic profiling, genomic data, and linked EHR (mean age at blood draw: 56 years, SD: 16 years; 55% female). Participants had extensive longitudinal clinical interactions with the MGB health system (median of 18 years and 238 visits; eFigure 1). Across the nine psychiatric conditions, prevalence ranged from 0.5% for ASD (N=116) and AN (N=137) to 40% for ANX (N=10,143), 33% for MDD (N=8,363), and 10% for SUD (N=2,598)

(eTable 2). 13,126 (52%). participants had at least one documented psychiatric diagnosis and 5,879 (23%) had at least two prescriptions for any psychotropic medication in the year prior to blood draw (eTable 3). In our sample, all PRS (except for ASD) were associated with their target disorders at a p-value < 0.05 (eFigure 2). PRS for AN, ANX, ADHD, ASD, and SUD explained 1% or less of variance in liability for their respective disorders, while PRS for SCZ, BD, PTSD, and MDD explained around 2% of variance in liability (eTable 1).

### PRS Associations Persist After Accounting for Diagnosis and Medication

We first examined the *total effect* of psychiatric genetic liability on circulating metabolites, and then assessed the attenuation after adjusting for lifetime diagnosis and psychotropic medication use (Figure 1). Across all PRS-metabolite pairs (eTable 5), we observed 604 significant associations at p< 1.36 ×10^-4^ in the *total effect model*. After adjustment for both diagnosis and medication use (*direct effect model*), all associations were directionally consistent, with only a small change in effect size (-10.3%, SD: 7%), and 536 (89%) remained significant (Figure 2). Attenuation was distributed across all metabolite categories eFigure 3. Requiring 2 phecodes to define cases resulted in similar effect size reduction (-9.7%, SD: 7%; 531 remained significant). In sensitivity analyses, restricting the sample to participants without psychiatric diagnoses or medications (N=11,150; 44%), effect sizes had a similar decrease but were more variable (-8.5%; SD: 19%), with 300 (50%) associations remaining significant, reflecting reduced statistical power due to the smaller sample size. After adjusting for BMI (N=21,305; 84%), associations remained directionally consistent but with a larger decrease in effect size, -39% (SD: 12%), with only 227 (38%) associations remaining significant.

**Figure 1.**
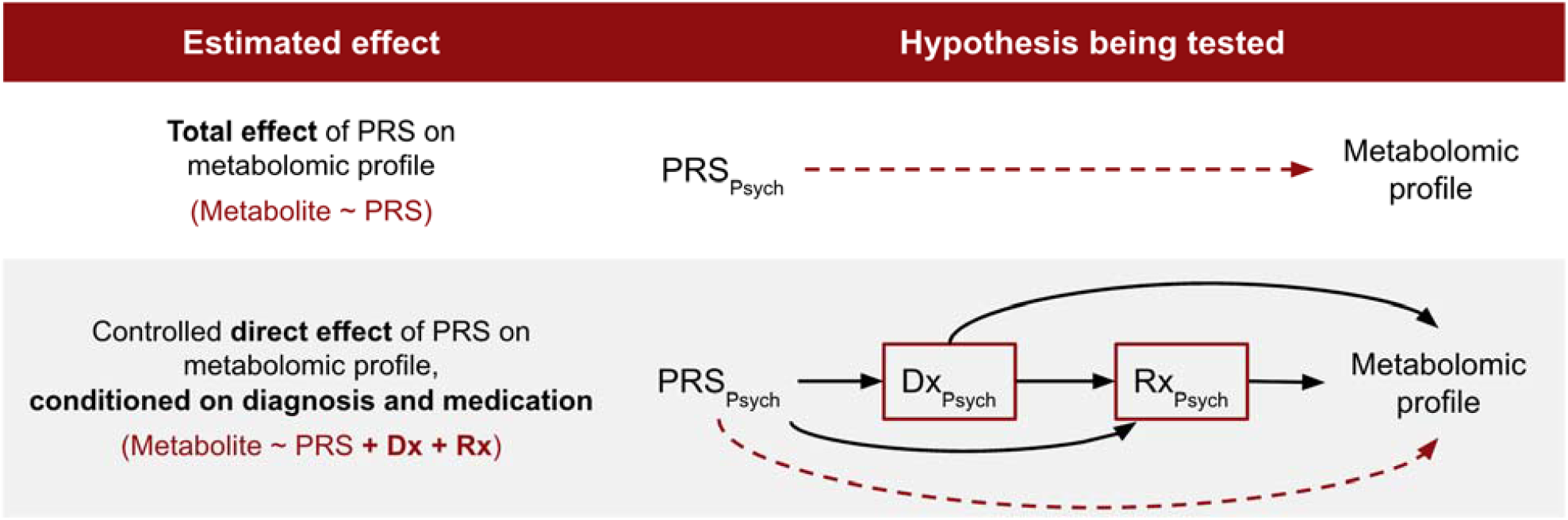
Causal diagram and study design. PRS_Psych_ denotes the polygenic risk score for a given psychiatric disorder; Dx_Psych_ denotes diagnosis of that disorder; Rx_Psych_ denotes psychotropic medication use. All models have been adjusted for age, sex assigned at birth, and top 10 principal components for population stratification. The dashed arrow from PRS_Psych_ to metabolite represents the direct effect being estimated. Genotype precedes illness onset and treatment, justifying the treatment of Dx_Psych_ and Rx_Psych_ as mediators of the PRS-metabolite association. *Direct model* tests whether the direct path persists after conditioning on diagnosis and treatment, assuming there is no measurement error or residual confounding. However, Dx_Psych_ and Rx_Psych_ may be imperfect proxies for illness and treatment effects, and residual confounding along these paths cannot be excluded.

**Figure 2.**
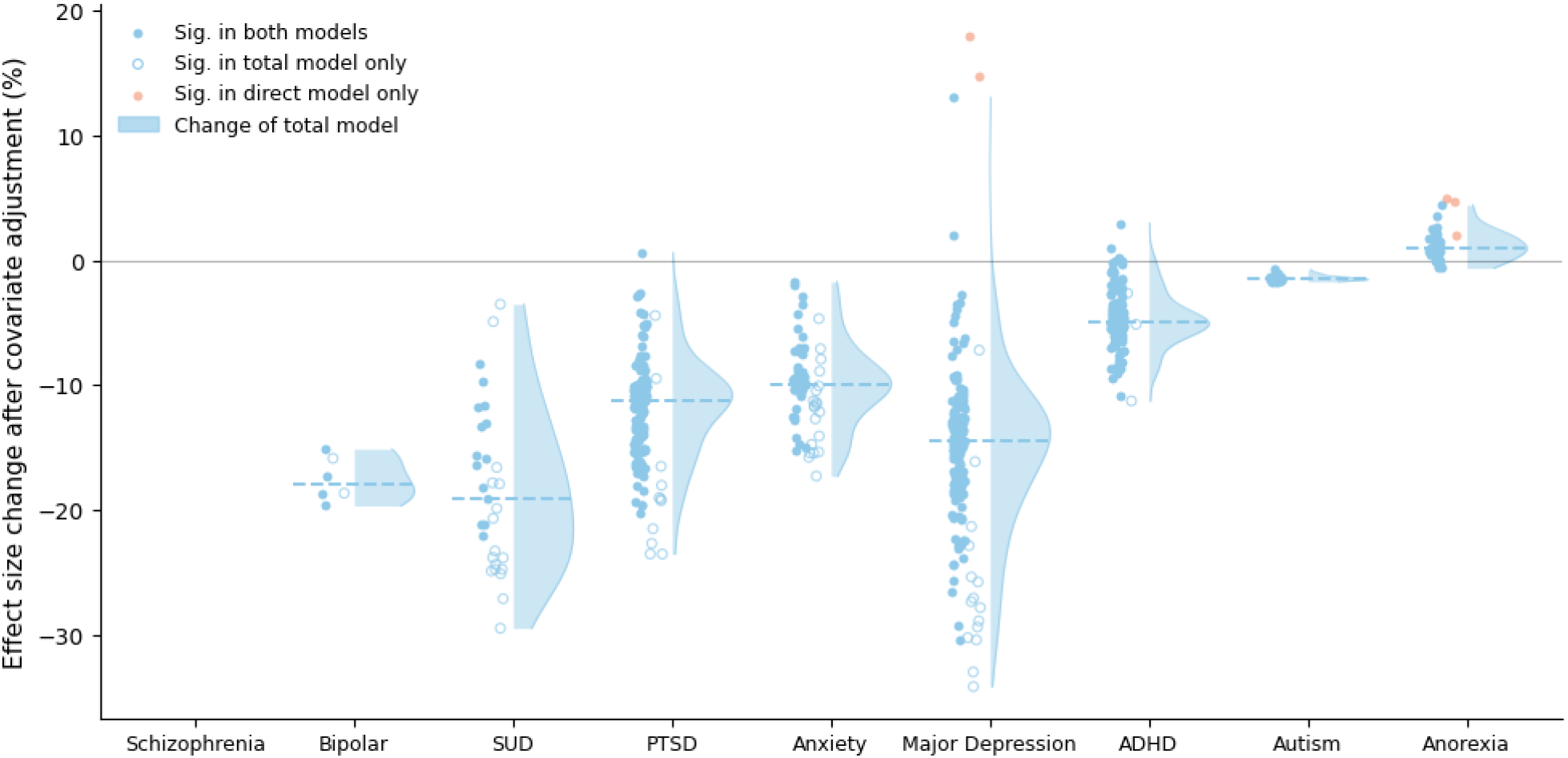
Change in PRS–metabolite associations after adjustment for lifetime psychiatric diagnosis and psychotropic medication use. For each PRS, the distribution (blue areas) shows the percentage change from the *total effect model* to the *direct effect model* ((β_direct_ – β_total_) / β_total_) × 100 across Bonferroni-significant associations (p< 1.36 ×10^-4^). The dashed line indicates the median. The horizontal solid line indicates no change. Across all PRS, the mean effect size change was -10.3% (SD: 7%), indicating minimal attenuation after covariate adjustment. AN was the only PRS showing a median change of +1%, reflecting a strengthening rather than attenuation of effects after adjustment.

PRS_SUD_ showed the largest attenuation after controlling for diagnosis and treatment (-18%, SD: 6%), whereas PRS_AN_ showed the largest attenuation after controlling for BMI (-48%, SD: 7%). In the most stringent sensitivity analysis, adjusting for diagnosis, treatment, and BMI, PRS_MDD_ retained 91 significant associations (the largest of all PRS tested) despite an average effect size change of -41% (SD: 11%). (eTable 5).

### Direct genetic effects on metabolite profiles

PRS_MDD_, PRS_ADHD_, and PRS_PTSD_ were associated with extensive metabolic dysregulation after accounting for diagnosis and treatment, with 142, 137, and 134 significant associations across the metabolome respectively (eTable 5). These three PRS had similar magnitude and direction of effects across lipid transport, inflammatory, and energy metabolism pathways (Figure 3). PRS_ANX_, PRS_ASD_, and PRS_SUD_ had fewer significant associations, with 50, 29, and 14 respectively. PRS_AN_ had 31 significant associations that were consistently in the opposite direction from PRS for other disorders, likely reflecting the negative genetic correlations between AN and adiposity, circulating lipids, and glycemic traits ^28^. Notably, PRS_SCZ_ and PRS_BD_ showed few associations (0 and 4 respectively), despite having the strongest association with their target diagnoses among all PRS tested (eFigure 2), suggesting that the well-documented cardiometabolic comorbidities in patients with SCZ and BD are unlikely to reflect a shared genetic etiology with cardiometabolic traits. While some associations were specific to a single condition, most metabolic signatures were transdiagnostic.

**Figure 3.**
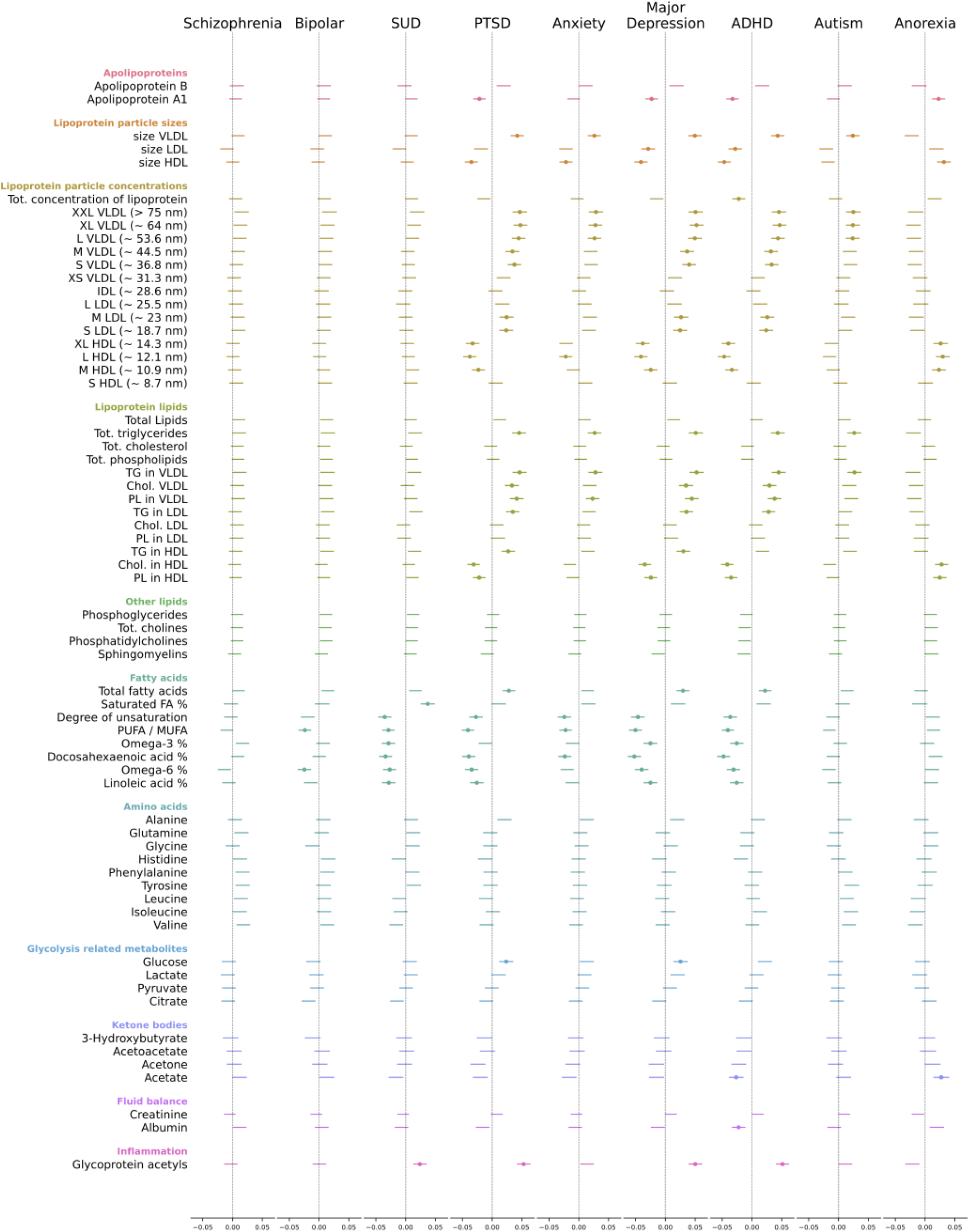
PRS associations with circulating metabolites after adjustment for lifetime psychiatric diagnosis and psychotropic medication use (*direct effect*). Each panel shows effect size estimates (β) and 95% confidence intervals for associations between a given PRS and 65 circulating metabolites grouped by molecular category. Point estimates are only shown for associations surviving Bonferroni correction (p < 1.36 ×10^-4^).

#### Dyslipidemia

PRS of internalizing (PTSD, ANX, MDD) and neurodevelopmental disorders (ADHD, ASD) were associated with elevated triglycerides across lipoprotein fractions (total triglycerides: β=0.026 to 0.051, all p<1.4 ×10^-5^; Figure 4A) and with increased VLDL particle size (β=0.024 to 0.049, all p< 2.6 ×10^-5^; Figure 4B). PRS_PTSD_, PRS_MDD_, and PRS_ADHD_ were additionally associated with decreased ApoA1, a marker of HDL concentration (β=-0.033 to -0.021, all p<6.3 ×10^-5^). This pattern of elevated triglycerides, large VLDL, and reduced HDL are associated with atherosclerosis ^29^, hypertension ^30^, insulin resistance ^31^, and metabolic syndrome ^32^.

**Figure 4.**
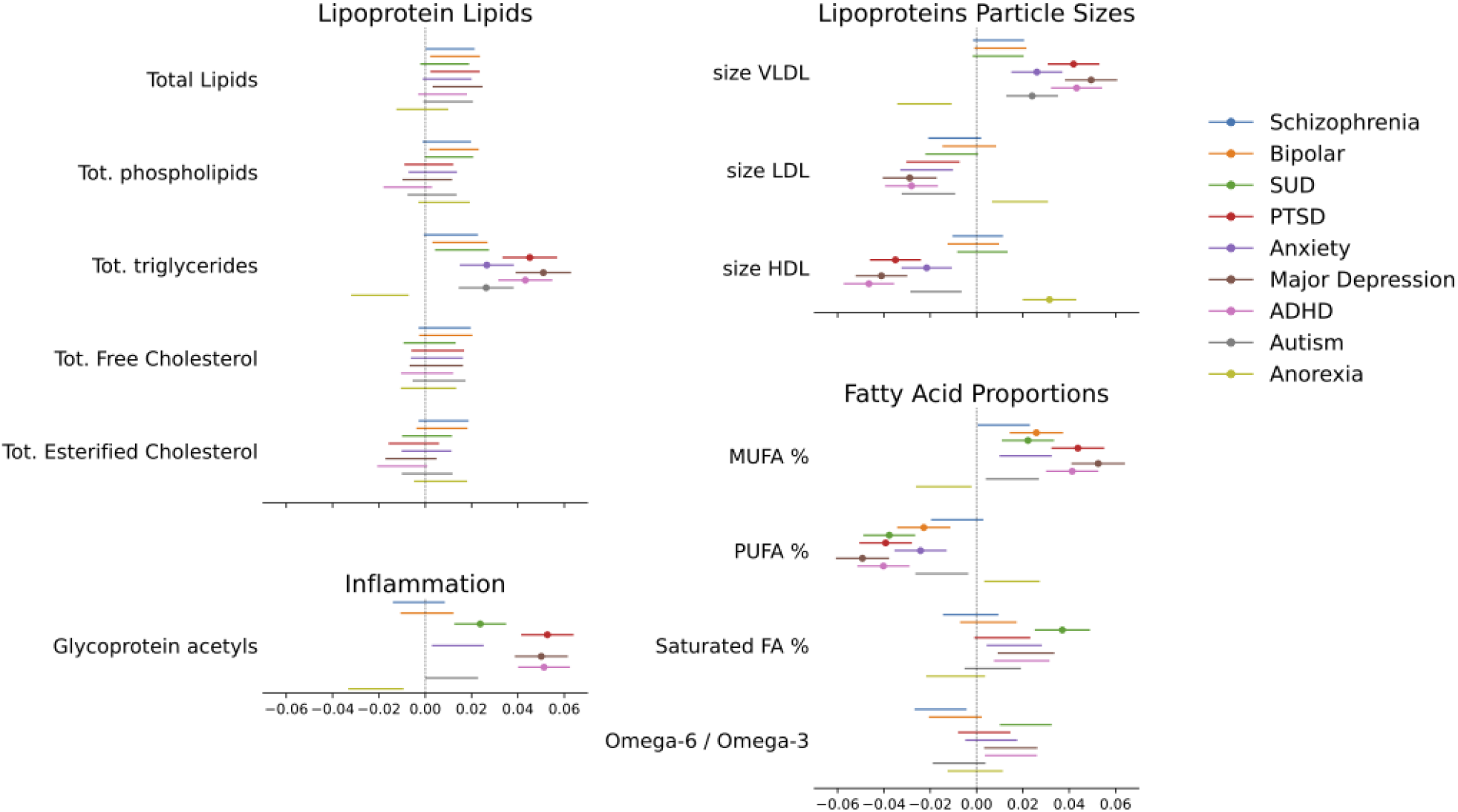
PRS associations with selected circulating metabolites after adjustment for lifetime psychiatric diagnosis and psychotropic medication use (*direct effect*). Horizontal lines represent 95% confidence intervals; dots indicate associations surviving Bonferroni correction (p < 1.36 ×10^-4^). Across internalizing and neurodevelopmental disorders, PRS were associated with elevated triglycerides and increased VLDL particle size but decreased HDL particle size. PRS for PTSD, MDD, ADHD, and SUD were additionally associated with elevated glycoprotein acetyls. Across disorders, fatty acid proportions shifted toward greater saturation (increased MUFA % and decreased PUFA %) with SUD characterized by the highest proportion of saturated fatty acids among all PRS.

#### Chronic inflammation

PRS associations with glycoprotein acetyls (GlycA) were among the strongest observed across all metabolites (Figure 4C, eTable 5). GlycA is a composite marker of chronic low-grade inflammation that, while correlated with CRP and IL-6 ^33^, is predictive of all-cause mortality independent of CRP ^34^. GlycA was associated with PRS_PTSD_, PRS_ADHD_, PRS_MDD_ (β=0.050-0.053, all p< 2.2 ×10^-16^), and with PRS_SUD_ (β=0.024, p= 3.2 ×10^-5^).

#### Fatty acid composition

PRS for BD, SUD, PTSD, ANX, MDD, and ADHD were all associated with a reduced ratio of polyunsaturated to monounsaturated fatty acids (PUFA/MUFA; β = −0.022 to −0.050, all p< 3.2 ×10^-5^), indicating a transdiagnostic decrease in unsaturated fatty acids, and consistent with prior evidence that PUFA supplementation may reduce the risk of various mental disorders psychiatric disorders ^35^. After BMI adjustment, associations persisted for all PRS (β = −0.022 to −0.037, all p< 3.6 ×10^-5^) except PRS_ANX_ (β = -0.015; p= 8.2 ×10^-3^). Additionally, PRS_SUD_ was uniquely associated with a higher percentage of saturated fatty acids (β=0.037, p=1.2 ×10^-9^; Figure 4D), a distinct signature consistent with prior findings in alcohol use disorder ^36,37^.

### Psychiatric genetic liability and aggregate cardiometabolic risk

To contextualize individual metabolite associations within clinically meaningful outcomes, we examined associations between psychiatric PRS and MRS for four cardiometabolic conditions (Figure 5). In the *direct effect model*, PRS_MDD_, PRS_PTSD_, and PRS_ADHD_ were each associated with increased MRS_T2D_ (β = 0.065 to 0.084, all p< 4.6 ×10^-6^), MRS_MI_ (β = 0.044 to 0.054, all p< 1.6 ×10^-8^), MRS_IS_ (β = 0.047 to 0.062, all p< 1.9 ×10^-6^), and MRS_VOD_ (β = 0.043 to 0.069, all p< 4.0 ×10^-8^). Notably, PRS_SUD_ showed a similar pattern of associations across MRS (β = 0.031 to 0.057, all p< 5.3 ×10^-5^), despite having fewer significant individual metabolite associations, suggesting that distinct genetic effects across metabolites can converge on similar cardiometabolic risk. PRS_ANX_ showed a more limited pattern, with associations restricted to MRS_T2D_ (β = 0.045, p=0.0013) and MRS_VOD_ (β = 0.025, p=0.0012), but not MRS_MI_ or MRS_IS_. PRS_BD_ was only associated with MRS_VOD_ (β=0.025, p=0.0012). PRS for SCZ, ASD, and AN showed no significant associations with any MRS (eTable 6).

**Figure 5.**
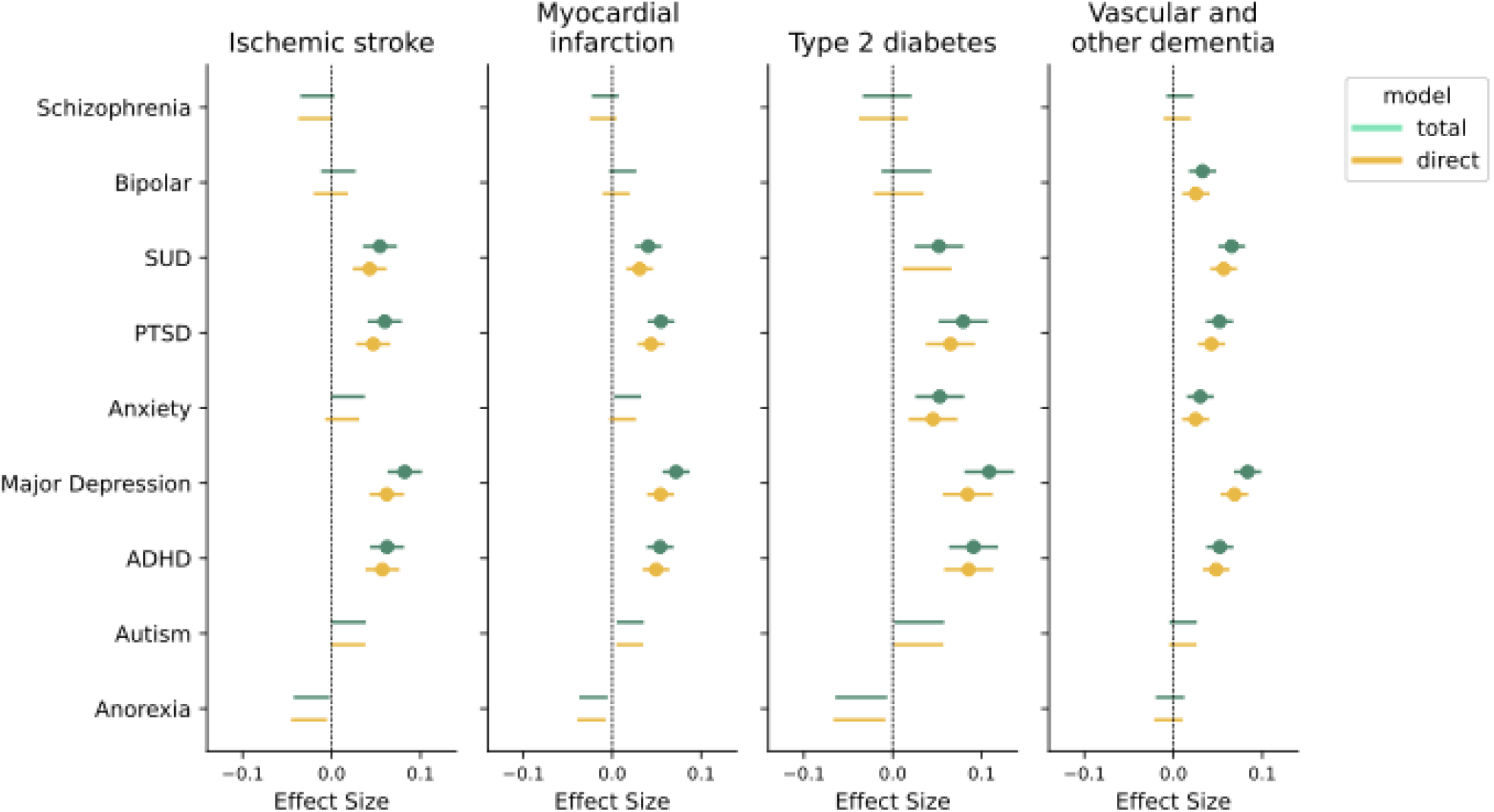
Associations between psychiatric PRS and metabolomic risk scores (MRS) for four cardiometabolic conditions, before and after adjustment for lifetime psychiatric diagnosis and psychotropic medication use. Each row represents a PRS predictor; each panel represents an MRS outcome. Horizontal lines represent 95% confidence intervals; points indicate associations surviving Bonferroni correction (p < 1.38 ×10^-3^). Total and direct effect estimates are shown in green and yellow, respectively.

Given the limited metabolite associations with PRS_SCZ_ and PRS_BD_ (despite the well-known comorbidity of SCZ and BD with cardiometabolic conditions), we examined the effect of diagnosis and medication use on cardiometabolic MRS in our fully-adjusted models. Notably, SCZ and BD diagnoses were associated with MRS_T2D_ (β=0.48 and 0.22, p=2.6×10^-4^ and 8.1×10^-4^, respectively) and MRS_VOD_ (β=0.25 and 0.18, p=6.1×10^-4^ and 2.1×10^-7^, respectively), after adjusting for their respective PRS and medication use. Additionally, antipsychotic medication use was associated with elevated MRS_MI_ (β=0.18, p=9.2×10^-4^) and MRS_IS_ (β=0.19, p=5.3×10^-3^), after adjusting adjusting for PRS_SCZ_ and diagnosis of SCZ.

## Discussion

In a sample of 25,290 individuals, we characterized the associations between polygenic risk for nine psychiatric disorders and 249 circulating metabolites and four cardiometabolic risk scores. We found that polygenic liability for MDD, PTSD, and ADHD is associated with a pattern of atherogenic dyslipidemia and systemic inflammation that is detectable in the absence of clinical illness and is independent of psychotropic medication use. PRS for these three disorders and PRS_SUD_ converged on elevated metabolomic risk scores for myocardial infarction, ischemic stroke, and vascular dementia, even though PRS_SUD_ had a distinct metabolomic profile characterized by elevated saturated fatty acids rather than altered lipoprotein composition. By contrast, PRS_SCZ_ and PRS_BD_ showed minimal effects on circulating metabolites despite these conditions being repeatedly linked to cardiometabolic disease in case-control studies, and despite these PRS explaining greater variance in their target diagnoses than other PRS examined. Lastly, PRS_AN_ was unique in that its effects were opposite in direction to those of other disorders and strengthened after adjustment for diagnosis and medication. While EHR-based measures capture an imperfect picture of illness burden and treatment exposure, and some residual confounding cannot be excluded, our findings support a partially pleiotropic model underlying cardiometabolic and psychiatric comorbidity, with genetic effects on peripheral metabolism exhibiting both shared and unique associations across psychiatric disorders.

Although internalizing and neurodevelopmental disorders differ in their clinical presentation and developmental course, their PRS converged on elevated triglycerides, recapitulating prior epidemiological ^10,38,39^ and genetic findings ^40,41^ linking each disorder independently to this trait. Similarly, a strong association with GlycA was shared across PRS_MDD_, PRS_PTSD_, and PRS_ADHD_. The combination of atherogenic dyslipidemia and elevated GlycA may point to a shared pathway driving both lipid dysregulation and chronic systemic inflammation ^42^, two processes that interact in the pathogenesis of T2D ^43^, and that have been proposed as defining a subtype of depression ^2^. Together, these findings extend the immuno-metabolic profile proposed for depression ^2^ to internalizing and neurodevelopmental disorders more broadly.

Interestingly, PRS_SCZ_ and PRS_BD_ showed minimal associations with peripheral metabolites, despite the well-known elevated risk of cardiometabolic disease among individuals with schizophrenia and bipolar disorder ^4,44^. Nevertheless, these results align with prior genetic correlation ^45^ and genetic epidemiologic studies ^13^. Our results therefore suggest that the elevated cardiometabolic risk in these disorders is less likely attributable to shared genetic etiology than to secondary effects of illness or treatment, such as antipsychotic-induced dysglycemia and dyslipidemia ^46^. Consistent with this interpretation, we found that SCZ and BD case status, as well as antipsychotic use, were each independently associated with cardiometabolic MRS.

BMI is genetically correlated with most psychiatric disorders ^8^ and is associated with circulating metabolite levels, which presents a challenge for determining whether BMI functions as a mediator or a potential source of collider stratification bias ^47^. In our sensitivity analysis, effect sizes decreased on average -39% after controlling for BMI, with the strongest attenuation observed for PRS_AN_ (-48%). Notably, PRS_MDD_ retained a large number of significant associations, suggesting its broad metabolic signature is only partially accounted for by BMI. This pattern supports a partial mediation model and aligns with previous findings where the association of psychiatric PRS with cardiometabolic disease was attenuated but not eliminated after adjustment for BMI and other lifestyle factors ^13^.

The association of PRS_MDD_, PRS_PTSD_, PRS_ADHD_, and PRS_SUD_ with elevated MRS across cardiometabolic conditions indicates that their individual metabolite associations aggregate into a pattern of clinically meaningful cardiometabolic dysregulation. Consistent with these results, Bergstedt et al. ^13^ showed that PRS_MDD_, PRS_PTSD_, and PRS_ADHD_ increased risk for cardiometabolic disease at the population level, and Lawrence et al. ^48^ demonstrated genome-wide genetic correlations between internalizing, neurodevelopmental, and substance use disorders and a broad range of physical illness domains. Importantly, our results suggest potential mechanisms for these associations by identifying specific metabolic pathways operating independent of psychiatric diagnosis and treatment.

Prior work describing genomic correlations between psychiatric disorders and cardiometabolic outcomes found a notable divergence between MDD and SCZ/BD, situating MDD closer to T2D in its genetic correlation with circulating metabolites ^45^. Our findings extend this work by contextualizing this divergence within a broader set of psychiatric disorders and by grounding it in the metabolomes of individual participants rather than genome-wide correlations. Future work should expand metabolic coverage through more comprehensive platforms, employ colocalization analyses to identify the specific genetic variants and pathways driving these associations, and leverage longitudinal designs to strengthen causal inference and examine gene-environment interactions.

### Limitations

Several limitations of our study should be considered. First, the interpretation of PRS-metabolite associations is complicated by potential bidirectional relationships between psychiatric and metabolic traits, supported by Mendelian randomization studies in both directions ^45,49,50^. The psychiatric GWAS used to construct these PRS did not restrict participation based on metabolic comorbidities, meaning the PRS may capture shared psychiatric-metabolic signals from the source population and inflate associations in the direction consistent with our findings. However, the magnitude of this bias is difficult to characterize. Second, psychiatric phenotyping relies entirely on ICD codes from the EHR, which may be prone to misclassification ^19^ and fail to capture illness severity or duration, making them an imperfect proxy for adjusting for illness effects. For example, a recent mild substance use disorder and a decade-long history of heavy alcohol use with secondary hepatic and metabolic consequences receive the same binary indicator. Third, participants of the MGB Biobank receive care at an academic medical center, which may introduce selection bias that is difficult to fully characterize ^51^. Relatedly, due to sample size constraints, all analyses were restricted to participants of European genetic ancestry, limiting generalizability of our findings to the broader population. Finally, all effect sizes are modest, reflecting associations at the population level and should not be interpreted as warranting clinical application as patient-level biomarkers.

## Conclusion

Polygenic liability for several psychiatric disorders is associated with specific patterns of metabolite dysregulation, independent of clinical illness and psychotropic medication use, supporting a model in which cardiometabolic comorbidity in psychiatric disorders partly reflects their pleiotropic overlap. The shared biological pathways identified here, particularly lipid dysregulation and chronic inflammation, may represent targets for interventions that simultaneously address psychiatric and cardiometabolic risk.

## Data Availability

The MGB Biobank is available to MGB researchers with appropriate IRB approval.

## Acknowledgments

JFDLH and TG are supported by R01MH130899 from the National Institute of Mental Health (NIMH). YHL is supported by the NARSAD Young Investigator Grant (33456) from the Brain & Behavior Research Foundation. JDT is supported by K01MH141330 from the NIMH. MC is supported by the Mass General Brigham Training Program in Precision and Genomic Medicine (T32HG010464) from the National Human Genome Research Institute. WUM is supported by the Biomedical Informatics and Data Science Research Training Program (T15LM007092) from the National Library of Medicine. YC is supported by K01HL179256 from the National Heart Lung and Blood Institute (NHLBI). JWS is supported by R01MH118233 and R01MH137218 from the NIMH. JL-S is supported by R01HL169300, 1OT2HL161841, and R01HL155742 from the NHLBI.

JWS is a member of the scientific advisory board of Sensorium Therapeutics (with stock options) and has received grant support from Biogen. JL-S is a scientific consultant to TruDiagnostic and Antipode.

This study would not be possible without the contributions of Mass General Brigham (MGB) patients and Biobank participants. We would also like to thank the research coordinators and the Biobank study for their tremendous effort in participant recruitment and sample collection. Lastly, we would like to acknowledge the RPDR team for their work maintaining the enterprise research patient data warehouse.

**Main Exhibits**

