## Supplementary Methods and Figures for "Association of Genetic Liability to Psychiatric Disorders with Peripheral Metabolic Dysregulation"

### Supplementary Materials

**___________**

**eMethods.** Genomic quality control (QC) procedure and Metabolomic QC procedure at MGB Biobank.

**___________**

**eFigure 1.** Distribution of healthcare utilization within MGBB.

**eFigure 2.** PRS performance in our sample across nine psychiatric conditions.

**eFigure 3.** Attenuation of PRS–metabolite associations by metabolite group.

**___________**

**eTable 1.** Source GWAS for PRS and effect sizes in our sample.

**eTable 2.** Crosswalk between Phecode and ICD codes for nine psychiatric disorders.

**eTable 3.** Medications used in analysis, with information on risk of metabolic side effects and number of individuals taking them in the year prior to blood draw

**eTable 4.** Nightingale NMR panel components.

**eTable 5.** Associations between PRS and metabolites without adjustment for diagnosis, medication, and BMI.

**eTable 6.** Associations between PRS and MRS with adjustment for diagnosis, medication, and BMI.

**This supplementary material has been provided by the authors to give readers additional information about their work.**

#### eMethods.

#### Genomic QC procedure at MGB Biobank.

The MGB Biobank utilized two Illumina genotyping arrays at different processing waves: the global screening array (GSA) and the multi-ethnic genotyping array (MEGA). QC and imputation were performed separately for each array, with some individuals overlapping. For all analyses described here, MEGA genotypes were retained for individuals who were genotyped using both arrays. Detailed information on the quality control and imputation procedures can be found at <https://github.com/Annefeng/PBK-QC-pipeline> and <https://github.com/getian107/MGBB-QC>. SNP-level QC included the removal of variants with high missingness, batch effects, evidence for deviation from Hardy-Weinberg equilibrium, or having a low imputation quality score. Individual-level QC included removal of samples which were outliers on autosomal heterozygosity, exhibited high missingness across SNPs, or which differed in their chromosomal and self-reported sex. Imputation was performed on the Michigan Imputation Server using the combined HRC/1000 Genomes reference panel. The current analyses were restricted to individuals with European-like genomes, identified using a random forest classifier trained on data from the 1000 Genomes Project (The 1000 Genomes Project Consortium 2015). Samples were further restricted to a maximally unrelated subset (identity-by-descent sharing < 0.2).

#### Nightingale Metabolomics, and the QC process at MGB Biobank.

The Nightingale panel includes measurements of 249 metabolomic biomarkers derived from nuclear magnetic resonance (NMR) spectroscopy. These biomarkers span key metabolic pathways, including lipids, amino acids, and glycolysis-related metabolites. A sample of Biobank plasma samples collected between May 2010 and August 2023 were sent to the Nightingale laboratory between October 2023 and March 2024. Nightingale uses a high-throughput NMR spectroscopy platform to quantify metabolic biomarkers from EDTA plasma samples. Samples are prepared by mixing plasma with phosphate buffer before being loaded into 96-well plates and analyzed using a Bruker AVANCE III HD 500 MHz spectrometer equipped with a nitrogen-cooled triple resonance probe. Multiple spectra are acquired per sample, including lipid and low-molecular-weight metabolite profiles, with quantification performed through proprietary deconvolution algorithms calibrated against internal and external standards (Würtz, 2017).

We applied QC steps following precedents in the UK Biobank (Julkunen, 2023; Ritchie 2023). The QC steps applied to the processed data are as follows:

1. Impute zero and missing (NA) values with half the minimum observed value for each biomarker
2. Exclude outliers beyond 4 times the interquartile range (IQR) from the median
3. Apply a natural log transformation
4. Use linear regression to adjust for spectrometer effects and extract residuals
5. Standardize the residuals
6. Individuals with more than 5 missing values were removed.

All preprocessing steps were implemented in R (version 4.1.2) using validated packages and custom scripts.

#### eFigure 1. Distribution of date and age at blood collection, and distribution of healthcare utilization (visits) at MGB within the analytic sample.

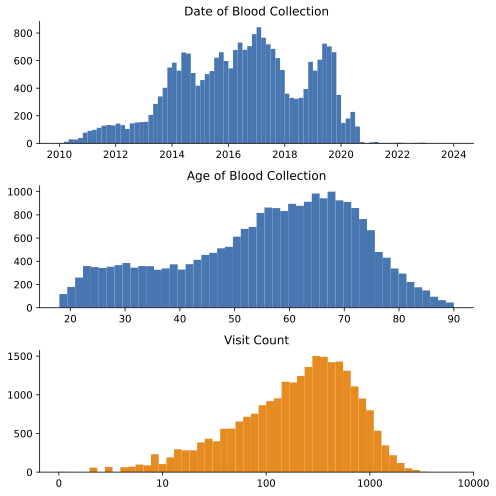

#### eFigure 2. PRS performance in our sample across nine psychiatric conditions.Variance explained on the liability scale (left panel; assumed population prevalences in eTable 1) and effect size per standard deviation increase in PRS (right panel). Error bars indicate 95% confidence intervals.

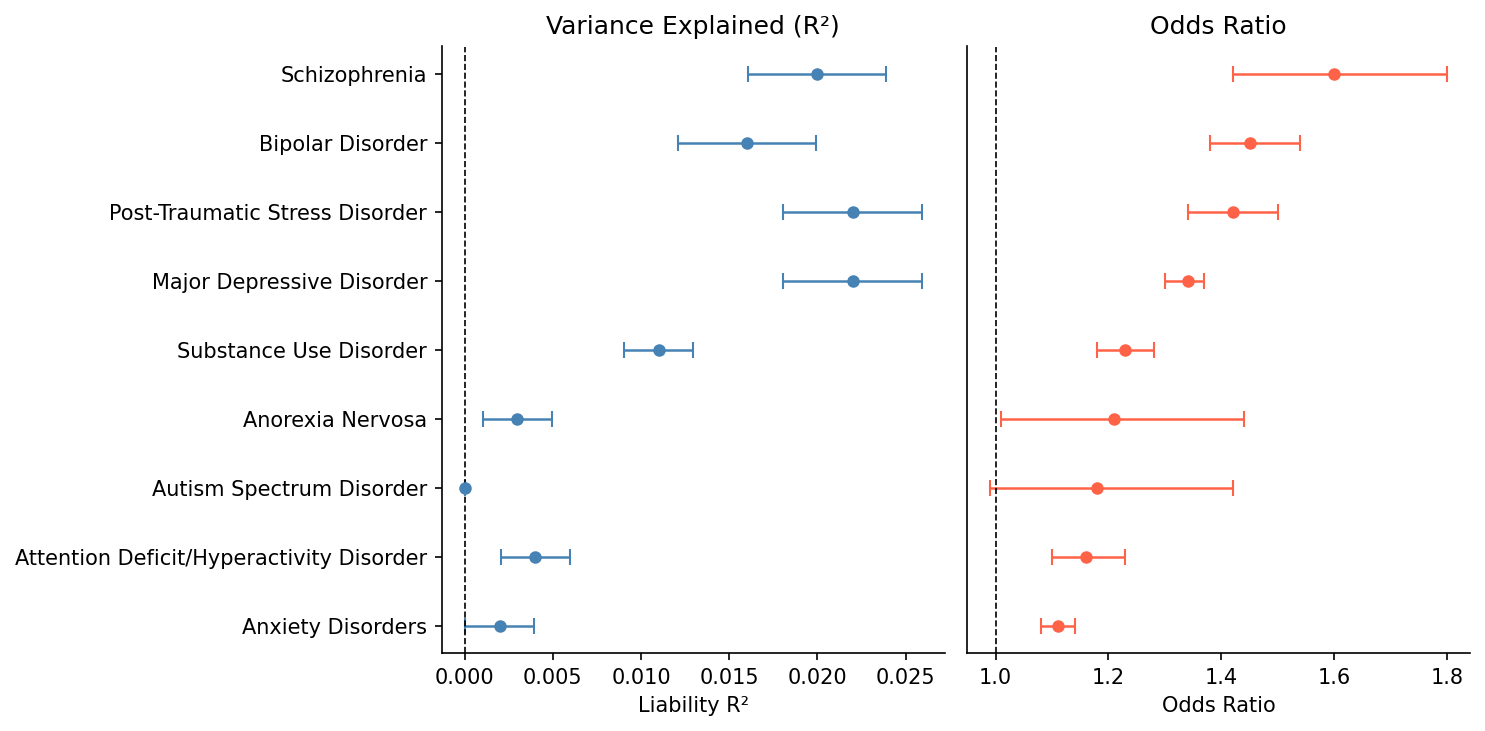

#### eFigure 3. Attenuation of all PRS–metabolite effect sizes after adjustment for lifetime psychiatric diagnosis and psychotropic medication use Effect attenuation is presented by metabolite category rather than by PRS for a given diagnosis (Figure 2). For each metabolite category, the distribution (blue) shows the percentage change in effect size from the *total effect model* to the *direct effect model* (( β_direct_ – β_total_ ) / β_total_ × 100) across Bonferroni-significant associations (p < 1.36 x10^-4^). Negative values indicate attenuation after covariate adjustment. The horizontal solid line (0%) indicates no change. The dashed line indicates the median for each category. Across all PRS, the mean effect size change was -10.3% (SD: 7%), indicating minimal attenuation after covariate adjustment. Attenuation was distributed across all metabolite categories, with no category showing markedly greater attenuation than others.

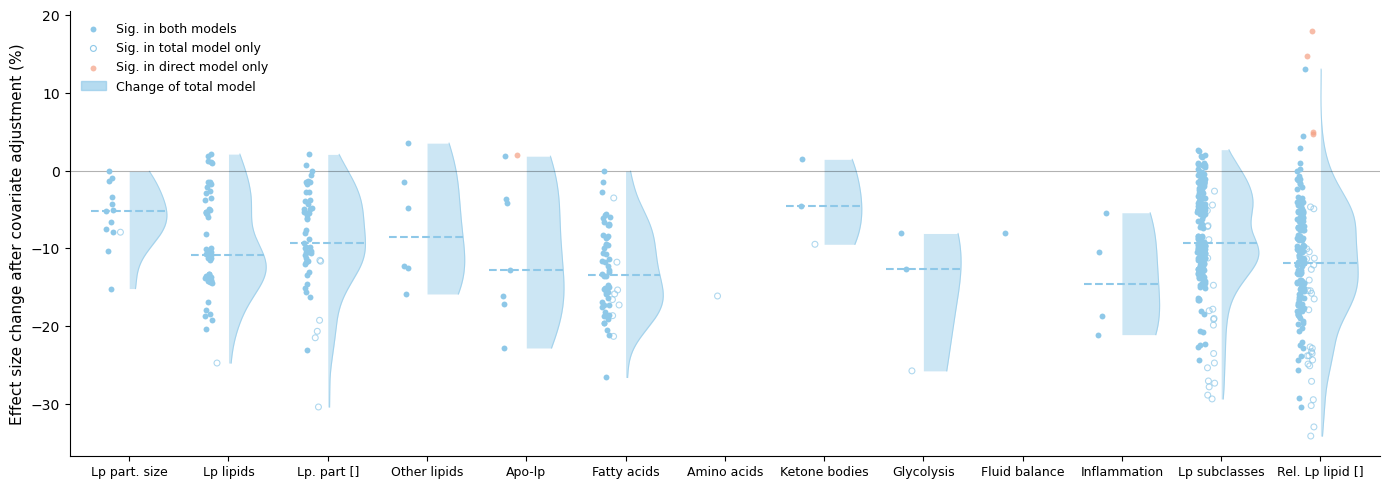
